# Chronotype and Cardiovascular Disease Risk Factors among Middle-aged and Older Adults: An observational and Mendelian Randomization Study

**DOI:** 10.64898/2026.01.12.26343964

**Authors:** Sina Kianersi, Kaitlin Potts, Heming Wang, Tamar Sofer, Raymond Noordam, Martin K Rutter, Kathryn Rexrode, Susan Redline, Tianyi Huang

## Abstract

**Introduction:** Circadian misalignment is an emerging risk factor for poor cardiovascular health, and chronotype may reflect underlying circadian processes. While previous conventional observational studies have reported adverse associations between evening chronotype and individual cardiovascular risk factors, Mendelian randomization (MR) may provide further insights into the role of chronotype in overall cardiovascular health, as measured by the American Heart Association’s Life’s Essential 8 (LE8; a composite lifestyle and cardiovascular health score ranging from 0 to 100; higher scores indicate better health).

**Methods:** We conducted both observational cross-sectional and one-sample MR analyses among 317,730 UK Biobank (UKB) participants of White ethnicity. Chronotype was self-reported and modeled on a five-level continuous scale from “definitely evening” to “definitely morning”. A polygenic risk score including 341 morning chronotype-associated SNPs from a UKB GWAS served as the MR instrument. Two-stage least-squares regression estimated difference in LE8 per one-unit increment in chronotype towards more morningness, adjusting for age, sex, assessment center, genotyping batch, and 40 genetic principal components. To mitigate potential winner’s curse bias in UKB due to inflated GWAS estimates, we replicated the analysis in 13,396 White women in the Nurses’ Health Study II (NHSII).

**Results:** In UKB, the multivariable-adjusted difference in LE8 score for each one-unit increment toward more morningness was 0.75-points higher (95% CI: 0.72, 0.78; P<0.001) in observational analysis and a 0.75-points higher (95% CI: 0.55, 0.96; P<0.001) in MR analysis. MR results were similar for men and women (P-heterogeneity = 0.70). In NHSII, while both estimates were positive, increased morningness was associated with higher overall LE8 scores in observational analysis (β = 1.57, 95% CI: 1.40, 1.75; P<0.001), but not in MR analysis (β = 0.89, 95% CI: - 0.67, 2.44; P = 0.26), although the MR association became significant when the score was based only on behavioral components (β = 2.04; 95% CI: 0.43, 3.65; p = 0.0130). Further, morning chronotype was consistently associated with healthy diet across observational and MR analyses in both cohorts.

**Conclusions:** Our findings suggest a modest causal relationship between morning chronotype and better cardiovascular health profiles, particularly diet quality, although replication in other populations remains necessary.

## INTRODUCTION

Cardiovascular disease (CVD) remains the leading cause of mortality and morbidity, yet it is largely preventable by modifiable health behaviors (e.g., smoking) and health factors (e.g., elevated blood pressure) (1, 2). To provide a comprehensive framework for promoting cardiovascular health, the American Heart Association (AHA) introduced Life’s Essential 8 (LE8) in 2022, a holistic metric that captures diet, physical activity, nicotine exposure, sleep health, body mass index (BMI), blood lipids, blood glucose, and blood pressure (2). The circadian system, comprising central and peripheral clocks, has been shown to play a role in influencing each of these LE8 components (3, 4). Blood pressure, glucose metabolism, caloric intake and rest-activity patterns all exhibit pronounced circadian rhythms (3, 5). Circadian misalignment, where external cues (e.g., light) and internal clocks become desynchronized, may disrupt these rhythms, contributing to poor cardiovascular health and increased CVD risk (3–6).

Chronotype, defined as an individual preference for earlier (morning) or later (evening) timing of the sleep–wake cycle, reflects underlying differences in circadian timing and alignment It is influenced by genetic, environmental, and sociodemographic factors (8–10). In middle-aged and older adults, roughly 8% to 11% report as evening chronotypes, 24% to 35% morning chronotypes, and the remainder an intermediate type (11, 12). Evening types are more susceptible to circadian misalignment because common work and social schedules often impose earlier activity times, whereas morning types tend to follow routines that align more closely with their endogenous clock, potentially supporting better cardiovascular health (13).

In a study among female nurses, we found that self-reported evening chronotype, compared to the morning type, is associated with an overall unhealthy lifestyle based on a composite score of smoking, sleep, physical activity, BMI, diet, and alcohol use (11). Similarly, among UK Biobank (UKB) participants, compared with intermediate chronotypes, self-reported definite evening types were more likely to have poor overall LE8 scores, whereas definite morning types were slightly less likely (12). Previous studies have also reported associations between evening chronotype and adverse individual cardiovascular risk factors, such as poor diet quality (11, 12), heavy alcohol consumption (14), smoking (15), dyslipidemia and hyperglycemia (16, 17). Nonetheless, most previous studies, including ours, had a cross-sectional design, limiting causal inference due to the inability to assess temporality and the potential for reverse causation and residual confounding. Therefore, a Mendelian randomization (MR) analysis could provide further evidence regarding the potential causal relationships between chronotype and cardiovascular health (8, 18, 19).

In this study, we aimed to use MR approach to evaluate the associations between chronotype and cardiovascular health, assessed by the overall LE8 score and its eight individual components. For comparison, we also examined the corresponding observational associations within the same study sample. We hypothesized that greater morning chronotype, compared to greater evening chronotype, would be associated with healthier cardiovascular profiles, as reflected by higher overall LE8 scores and more favorable individual LE8 components, consistent with observational findings.

## METHODS

### Study design, settings, and participants

We conducted cross-sectional and individual-level one-sample MR analyses separately in the UKB (2006–2010) and the Nurses’ Health Study II (NHSII; 2009). The UKB is a large-scale population-based cohort initiated in 2006, including approximately 502,650 UK participants aged 40 to 69 years at recruitment (20). Invitations were mailed to nine million adults in England, Scotland, and Wales, identified through UK National Health Service patient registers (covering ∼98% of the population), yielding a response rate of ∼5.5% (20). As of March 2025, data release version 19.1, it includes 501,978 participants. From 2006 to 2010, baseline data were collected at 22 centers where participants provided electronic consent, completed questionnaires and interviews, underwent physical assessments, and gave blood and urine samples. Among the 501,936 participants, we excluded 6,170 with missing chronotype data (exposure), 161,722 with incomplete data for one or more LE8 components (primarily physical activity), 13,996 who self-identified as non-White, and 2,318 with no or low-quality genetic data. The final analytical sample comprised 317,730 participants (Supplemental Figure 1.A). UKB received ethical approval from the National Health Service North-West Multi-Centre Research Ethics Committee, and all participants provided written informed consent at enrollment.

The NHSII cohort, initiated in 1989, recruited 116,429 female registered nurses aged 25–42 years (90% White) by mail from nursing-board registries in 14 U.S. states, achieving a 24% response rate (21). At enrollment and every two years thereafter, participants completed comprehensive questionnaires covering lifestyle and health factors, with follow-up response rates consistently above 85% (21). From 1996-99, a subset of participants provided blood samples for genotyping. Among 15,219 participants who returned the 2009 questionnaire, when chronotype was assessed, and who also had genotype data, we excluded those who self-identified as non-White (n = 777), had missing chronotype data (n = 773), or were missing any LE8 components (n = 273), resulting in an analytical sample of 13,396 (Supplemental Figure 1.B). The NHSII study protocol received approval from the Institutional Review Boards of both Brigham and Women’s Hospital and the Harvard T.H. Chan School of Public Health. We followed STROBE and STROBE-MR guidelines to report our findings (22, 23).

### Self-reported chronotype

In both cohorts, chronotype was assessed via self-report using a validated single-item question from the Morningness–Eveningness Questionnaire (MEQ; 2006-2010 in UKB and 2009 in NHSII) (7, 24). Response was coded as a continuous variable with the following values: −2 (definitely evening), −1 (more evening), 0 (intermediate based on “Do not know in UKB/Neither in NHSII”), 1 (more morning), and 2 (definitely morning); “Prefer not to answer” was set to missing. The same coding approach has been previously used in genome-wide association studies (GWAS) of chronotype (8, 18). Of note, previous studies in UKB have also treated the “Do not know” response as an intermediate chronotype (8). The single-item chronotype measure shows strong correlation with the full MEQ score (25). Chronotype demonstrated moderate within-individual stability over longitudinal follow-up in both cohorts, with a kappa agreement of 0.63 over 6 years in NHSII and 0.72 over a mean of 10 years in UKB (11, 12).

### Genetic instrument for chronotype

In UKB, baseline blood-derived DNA was genotyped on two closely related Affymetrix arrays, the UK BiLEVE Axiom and the UKB Axiom, directly genotyping 805,426 variants (8, 26). Centrally applied quality control excluded samples with >5% missing genotypes, Hardy–Weinberg-equilibrium (HWE) departures, or excess heterozygosity, and variants with >5% missingness (26). Imputation against the Haplotype Reference Consortium (HRC) and UK10K + 1000 genome reference panels expanded genomic coverage to ∼96 million variants. Imputed genotypes were aligned to the positive strand of the reference genome, with variant positions reported according to GRCh37 coordinates (Data-Field 22828, version Aug 2025). Genotyping in NHSII was conducted using mainly blood samples and four high-density Illumina arrays: HumanCoreExome (n = 7,871), OncoArray (n = 2,990), Global Screening Array (n = 1,813), and HumanHap (n = 722). After standard quality control (removing variants with HWE violations, call rate <95%, imputation r^2^ < 0.30, or minor allele frequency, MAF, <0.01), genotypes were phased and imputed to the 1000 Genomes Project Phase 1 Integrated Release v3 reference panel (hg19/GRCh37), yielding ∼31 million variant (27).

The chronotype genetic instrument comprised 341 independent genome-wide significant (P <5×10^-8^) SNPs identified using BOLT-LMM, which accounts for population structure and relatedness, in a sex-combined UKB GWAS of 449,734 White European participants, with adjustment for age, sex, study center, and genotyping release (8). In the GWAS, chronotype was coded as a continuous variable ranging from −2 to 2, consistent with the approach described above (See Self-reported chronotype). Importantly, the identified SNPs were also validated against accelerometer-derived measures (8). Female-specific effect estimates for the same loci were obtained from a separate GWAS among UKB female participants using the same pipeline (GWAS sample size = 244,207) (18). The estimated heritability of chronotype was 13.7% (8). The GWAS quality control excluded variants with HWE P <1×10^-12^, imputation r^2^ <0.30, or MAF ≤ 0.1%. The GWAS was restricted to ∼12 million variants imputed against the HRC panel Across the 341 chronotype SNPs, the UKB imputation INFO scores, which reflect imputation quality, were excellent (mean = 0.99; interquartile range = 0.01). In NHSII, the corresponding mean for imputation accuracy (r^2^) across the 341 SNPs was 0.96 (interquartile range = 0.04). In UKB, the minimum MAF among the 341 SNPs was 1.17%. This was 1.04% in NHSII.

Following established guidelines (28), we built a chronotype polygenic risk score (PRS) from the 341 SNPs. In UKB, where the discovery GWAS and study sample overlap, similar to a previous study (18), we summed the imputed dosages of morningness-increasing alleles without weighting to minimize winner’s-curse/weak-instrument bias. In NHSII, an independent cohort, we applied female-specific β-coefficients from the UKB female GWAS to the 341 SNPs as weights. Of note, in NHSII, one SNP at chr3:8817423 (rs149611468) had an MAF below 1% only in the Global Screening Array; therefore, the PRS for 1,813 NHSII participants was derived from 340 SNPs. In both cohorts, scores were calculated from imputed dosages, preserving imputation uncertainty, with all variants aligned to the positive GRCh37 strand; palindromic SNPs with a MAF >42% were retained in main analyses (16 in NHSII; not applicable to UKB as it is the discovery GWAS dataset (8)).

### Life’s Essential 8

The LE8 framework includes eight components: four health behaviors (diet, physical activity, nicotine exposure, and sleep health) and four health factors (body mass index [BMI], blood lipids, blood glucose, and blood pressure). Following AHA guidelines, we assigned each LE8 component a 0–100 score, with higher values reflecting better cardiovascular health. We then averaged the eight component scores (unweighted) to produce an overall LE8 score on the same 0–100 scale (2). All four health behaviors were self-reported in both cohorts. In UKB, all four health factors were objectively assessed. In NHSII, BMI and blood pressure were self-reported, while data on blood lipids and blood glucose were unavailable. Hence, the overall score in NHSII is based on the six measured components. A validation study showed that scores derived from a subset of the eight components still reliably reflect overall cardiovascular health (29). Further details on data sources, assessment methods, as well as evidence for their validity, are provided in the Supplementary Methods and Supplemental Table 1.

### Confounders

In UKB, baseline sociodemographic data (2006–2010) were collected via the touchscreen questionnaire, capturing age, sex, ethnicity, educational attainment, family history of cardiovascular disease, and employment/shift-work status. Responses of “Do not know” or “Prefer not to answer” were coded as missing. Employment/shift-work classification was based on paid work status and the frequency of hours worked outside the conventional 9am-5pm schedule; individuals not in paid employment were assigned to a separate category. Area-level deprivation was quantified with the Townsend Deprivation Index, calculated from the most recent national census output-area data. In NHSII, age in 2009, census tract household income, geographic region, population density, menopausal status, postmenopausal hormone therapy, cumulative months of rotating night shift work (at least 3 nights/month), any rotating night shift work within the past two years, and family history of CVD were used as covariates.

### Statistical methods

We reported means (SD) for continuous variables and frequencies (%) for categorical variables. In cross-sectional analyses, we used separate multivariable linear regression models to assess associations between self-reported chronotype and the overall LE8 score and its eight components. Models were adjusted for the measured confounders described above. Robust standard errors were used to ensure valid confidence intervals in the presence of unequal error variances, and results were reported as adjusted β-coefficients, representing mean difference in LE8, with 95% confidence intervals and two-sided p values.

In the one-sample MR analyses, we estimated the causal effect of chronotype on the overall LE8 score and its eight components using two-stage least-squares (2SLS) regression, with the chronotype PRS as the genetic instrument (unweighted PRS in UKB and weighted PRS in NHSII). In the first stage, we regressed self-reported chronotype (included as a continuous variable) on the PRS and prespecified covariates: age, sex, assessment center, the first 40 ancestry principal components (PCs), and genotype batch in UKB; and age, the first four PCs (following previous NHSII studies), and genotyping arrays in NHSII. In the second stage, we regressed each continuous LE8 outcome on predicted chronotype values obtained from the first stage, adjusting for the same covariates in each cohort, and obtained robust (heteroskedasticity-consistent) standard errors. β-coefficients from the second stage were interpreted as difference in the LE8 score per one-unit increase in chronotype towards morningness (range: –2 to 2), with 95% confidence intervals and two-sided p values.

To test the MR assumptions and assess whether the genetic instrument was strongly associated with chronotype (relevance), unrelated to confounders (independence), and affected LE8 outcomes only via chronotype (exclusion restriction), we performed multiple sensitivity analyses. We reported the F-statistic and R^2^ to assess whether the genetic instrument was strongly associated with chronotype. To assess whether the genetic instrument was unrelated to potential confounders, we examined associations between chronotype PRS and measured confounders. As most two-sample MR estimators, except MR-Egger, are also valid for one-sample MR (30), we applied fixed- and random-effect inverse-variance weighting (IVW), the weighted median (31), and the weighted mode (32) approaches to assess whether the PRS associated with LE8 outcomes only via chronotype (exclusion restriction) (19). Additionally, because the weighted mode estimate was sensitive to the choice of bandwidth parameter (phi), we reported results using both the default value (phi = 1) and a wider bandwidth (phi = 2). Cochran’s Q and I^2^ were also calculated to assess heterogeneity among SNP effects. A fourth assumption in our study is the homogeneity assumption, which requires that the effect of chronotype on LE8 is similar across individuals. We explored its plausibility through sex-stratified MR, and additional confounder adjustment (Townsend deprivation index, education, family history of CVD, and employment/shift work).

We performed additional analyses to test the robustness of our findings. In both cohorts, we excluded participants with a history of myocardial infarction (MI) and/or stroke at or prior to chronotype assessment to evaluate the association in healthy participants. In UKB, mirroring the approach of a prior study, we conducted a sensitivity analysis restricting the unweighted chronotype PRS to SNPs replicated at Bonferroni significance in the independent 23andMe cohort (23andMe GWAS sample size = 240,098; SNPs = 242) (8, 18).

In MR analyses for NHSII, we conducted the following sensitivity analyses: we excluded participants who reported any night shift work in the past two years to reduce potential misclassification of chronotype due to externally imposed sleep-wake schedules (excluded n = 1,678). We excluded palindromic SNPs with ambiguous strand orientation (A/T or C/G alleles and MAF> 0.42, 16 SNPs) and repeated the one-sample MR analysis. Furthermore, as self-reported blood pressure and body weight in NHSII are more prone to errors, we restricted the overall score to the four behavioral components (diet, physical activity, nicotine exposure, and sleep health) in a post-hoc one-sample MR analysis. Finally, we repeated the MR analysis using an LE8 diet score calculated with the Dietary Approaches to Stop Hypertension (DASH) score instead of the AHEI-2010 used in the main analyses.

To evaluate potential differences between UKB and NHSII, cohort-specific estimates from both observational and MR analyses were pooled using an inverse-variance weighted random-effect model (SAS %METAANAL macro (33)) and assessed heterogeneity with Cochran’s Q. We extracted genetic data using QCTOOL and bcftools and generated the chronotype PRS with PLINK v2.0. Cross-sectional analyses (stats package, lm) and one-sample MR (AER package, ivreg) were conducted in R. Python 3.10 was used for data visualization and data processing. UKB analyses were executed on the UKB Research Analysis Platform, and NHSII analyses on the Channing Division of Network Medicine computing cluster at Brigham and Women’s Hospital. Code is available from the first author upon request. Access to both datasets and computing platforms requires application and approval.

## RESULTS

### Baseline characteristics

The demographic characteristics of UKB and NHSII are shown in (Table 1, Supplemental Table 2, and Supplemental Table 3). Mean (SD) age was 57 (8) years (52% female) and 55 (4) years (100% female), respectively. Mean (SD) Townsend deprivation index was –1.6 (2.9) in UKB, indicating participants were less deprived than the national average. In NHSII, the census-tract household income in 2009 was $81,862 ($31,690), higher than the U.S. median household income at that time. “More morning” chronotype was most common (33%) in the UKB, whereas “definitely morning” dominated (34%) in NHSII; “definitely evening” was reported by 8% and 12%, respectively. Mean (SD) overall LE8 scores were 67 (12) in UKB and 71 (15) in NHSII. LE8 components were broadly similar across the cohorts; diet quality was higher among female UKB participants, whereas blood-pressure scores were better in NHSII.

**Table 1.** Participants Chronotype and Life’s Essential 8 Scores Stratified by Cohort.

|  |  |  |  |  |
| --- | --- | --- | --- | --- |
|  | UKB |  |  | NHSII § |
|  | Total<br>N = 317,730 | Female<br>N = 165,193 | Male<br>N = 152,537 | Total<br>N = 13,396 |
| Age (years), Mean (SD) | 56.87 (8.06) | 56.51 (7.99) | 57.26 (8.13) | 55.4 (4.4) |
| Socioeconomic status, Mean (SD) * | -1.58 (2.92) | -1.58 (2.88) | -1.58 (2.96) | 81862 (31690) |
| Shift work, n (%) † | 27712 (8.7%) | 11945 (7.2%) | 15767 (10.3%) | 9637 (72.0) |
| Chronotype, n (%) |  |  |  |  |
| Definitely evening | 26116 (8.2%) | 13316 (8.1%) | 12800 (8.4%) | 1654 (12.3) |
| More evening | 81598 (25.7%) | 42816 (25.9%) | 38782 (25.4%) | 2991 (22.3) |
| Intermediate | 28384 (8.9%) | 11246 (6.8%) | 17138 (11.2%) | 653 (4.9) |
| More morning | 105160 (33.1%) | 56346 (34.1%) | 48814 (32.0%) | 3519 (26.3) |
| Definitely morning | 76472 (24.1%) | 41469 (25.1%) | 35003 (22.9%) | 4579 (34.2) |
| Life's Essential 8 components ‡ |  |  |  |  |
| Diet score, Mean (SD) | 53.45 (31.62) | 60.51 (30.15) | 45.81 (31.40) | 39.8 (31.3) |
| Physical activity score, Mean (SD) | 72.15 (27.88) | 72.09 (27.67) | 72.21 (28.10) | 76.9 (34.2) |
| Nicotine exposure score, Mean (SD) | 71.86 (30.19) | 73.73 (29.46) | 69.83 (30.84) | 87.0 (23.9) |
| Sleep health score, Mean (SD) | 89.94 (18.00) | 89.99 (18.09) | 89.88 (17.89) | 89.7 (17.9) |
| Body weight score, Mean (SD) | 69.99 (28.09) | 72.83 (29.07) | 66.90 (26.65) | 69.3 (32.5) |
| Blood lipids score, Mean (SD) | 47.89 (29.06) | 48.47 (29.68) | 47.27 (28.37) | Not Available |
| Blood glucose score, Mean (SD) | 90.52 (19.59) | 91.60 (18.09) | 89.35 (21.02) | Not Available |
| Blood pressure score, Mean (SD) | 41.95 (32.09) | 48.04 (33.75) | 35.34 (28.76) | 60.1 (29.7) |
| Life's Essential 8 overall score | 67.22 (11.78) | 69.66 (11.76) | 64.58 (11.22) | 70.5 (15.4) |
| * Socioeconomic status was measured with Townsend Deprivation Index in UKB, with higher values indicating lower socioeconomic status. In NHSII, census tract household income (USD) is reported.<br>† In UKB, participants who were employed and reported sometimes/usually/always shift work are included. In NHSII, participants who reported engaging in night shift work (11 pm–7 am) in the last two years (2007-2009) are included.<br>‡ Life's Essential 8 components were scored as points, with higher values reflecting closer adherence to recommended cardiovascular health behaviors and factors. All NHSII participants were female. Blood glucose and blood lipid scores are only not available for NHSII sample.<br>§ All NHSII participants are female nurses.<br>UKB: Uk Biobank; NHSII: Nurses' Health Study II |  |  |  |  |

### Cross-sectional analyses

Among the 317,730 UKB participants, each one-unit increase toward greater morningness on the five-level chronotype measure was associated with a 0.75-points higher overall LE8 score (95% CI: 0.72, 0.78) after adjustment for sociodemographic characteristics, family history of CVD, and employment/shift-work status (Table 2). Among the 13,396 NHSII participants, each one-unit increase in chronotype toward greater morningness (on the five-level measure) was associated with a 1.57-points higher overall LE8 score (95% CI: 1.40, 1.75), after adjusting for sociodemographic characteristics, menopausal and hormone therapy status, shift work history, and family history of CVD. While the direction of the associations was consistent between the two cohorts, the meta-analysis suggested that the magnitude of the association was statistically larger in NHSII than UKB (pooled β = 1.16; 95% CI: 0.35, 1.97; p for heterogeneity <0.001; Table 2, Figure 1). In both cohorts, self-reported chronotype was associated with all LE8 components except blood pressure. Of note, greater morningness was associated with slightly poorer sleep health score in UKB but significantly better sleep health score in NHSII. The strongest association was for nicotine exposure score, followed by diet and physical activity scores in UKB. By contrast, the strongest association was observed for physical activity followed by body weight and diet scores in NHSII.

**Figure 1.**
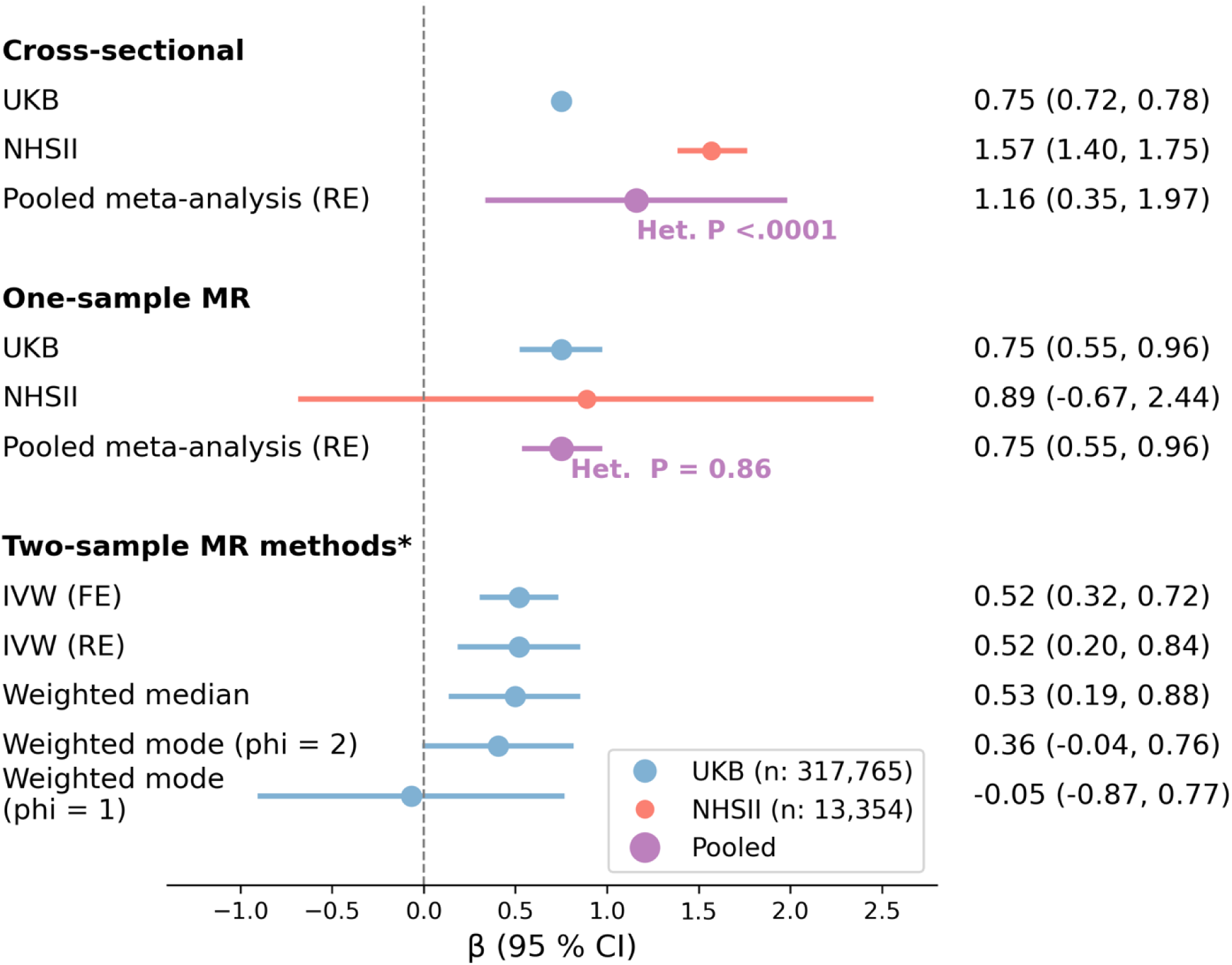
Forest Plot of the Associations Between Chronotype and Overall Life’s Essential 8 Scores

**Table 2.** Cross-Sectional Associations Between Chronotype and Life’s Essential 8 (LE8) Scores.

|  |  |  |  |  |  |  |  |  |
| --- | --- | --- | --- | --- | --- | --- | --- | --- |
|  | UK Biobank, N = 317,730 |  |  |  | Nurses' Health Study II, N = 13,396 |  |  |  |
| LE8 scores (range 0-100) | Beta (95% CI) Model 1* | P trend | Beta (95% CI) Model 2† | P trend | Beta (95% CI) Model 1‡ | P trend | Beta (95% CI) Model 2§ | P trend |
| Diet | **1.63 (1.55, 1.71)** | <0.001 | **1.30 (1.22, 1.38)** | <0.001 | **1.68 (1.33, 2.03)** | <0.001 | **0.92 (0.58, 1.27)** | <0.001 |
| Physical activity | **1.52 (1.44, 1.59)** | <0.001 | **1.28 (1.20, 1.35)** | <0.001 | **2.70 (2.29, 3.11)** | <0.001 | **1.56 (1.17, 1.95)** | <0.001 |
| Nicotine exposure | **1.86 (1.78, 1.94)** | <0.001 | **1.72 (1.64, 1.80)** | <0.001 | **0.73 (0.44, 1.03)** | <0.001 | **0.58 (0.28, 0.88)** | <0.001 |
| Sleep health | **-0.06 (-0.11, -0.01)** | 0.0182 | **-0.15 (-0.20, -0.10)** | <0.001 | **0.77 (0.55, 0.99)** | <0.001 | **0.60 (0.38, 0.82)** | <0.001 |
| Body weight | **0.42 (0.35, 0.50)** | <0.001 | **0.16 (0.09, 0.23)** | <0.001 | **2.37 (2.00, 2.75)** | <0.001 | **1.33 (0.98, 1.67)** | <0.001 |
| Blood lipid | **0.34 (0.26, 0.42)** | <0.001 | **0.18 (0.11, 0.26)** | <0.001 | Not available | - | Not available | - |
| Blood glucose | **0.23 (0.18, 0.28)** | <0.001 | **0.06 (0.01, 0.11)** | 0.0191 | Not available | - | Not available | - |
| Blood pressure | 0.05 (-0.03, 0.13) | 0.20 | -0.02 (-0.09, 0.06) | 0.69 | **1.19 (0.85, 1.53)** | <0.001 | 0.22 (-0.10, 0.55) | 0.17 |
| Overall score | **0.75 (0.72, 0.78)** | <0.001 | NA | NA | **1.57 (1.40, 1.75)** | <0.001 | NA | NA |
| - Results from linear regression models. Chronotype was treated as a continuous variable ranging from -2 (definitely evening) to 2 (definitely morning). LE8 scores were continuous dependent variables ranging from 0 to 100. *P*-trend was calculated by modeling chronotype as a continuous variable ranging from -2 to 2. Beta coefficients represent changes in LE8 score, in points, for each one-unit increase in the chronotype scale.<br>* Model 1 (UKB): adjusted for age, sex (female, male), assessment center, Townsend deprivation index, education, family history of CVD (yes, no), and employment/shift work (employed, never/rarely shift work; employed, sometimes/usually/always shift work; retired/Other).<br>† Model 2 (UKB): adjusted for Model 1 + the other seven LE8 components.<br>‡ Model 1 (NHSII): adjusted for age, census tract household income (U.S. dollars), region (Northeast, Midwest, South, West), population density per square kilometer, postmenopausal (yes, no), and postmenopausal hormone use (current, past, never), cumulative months of rotating night shift work (at least 3 nights/month), any night shift work (11 pm-7 am) in the previous 2 years (yes or no), and family history of CVD (yes or no).<br>§ Model 2 (NHSII): Adjusted for Model 1 + the other five LE8 components. |  |  |  |  |  |  |  |  |

### One-sample MR analysis

Across cohorts and weighting methods, the PRS for chronotype showed strong instrument strength in UKB (F statistics: 4961 - 7744; R^2^: 1.54% - 2.38%) and moderate strength in NHSII (F statistics: 174 - 179; R^2^: 1.28% - 1.32%), with all first-stage regression p values ≤1.91×10^-39^ (Table 3). The weighted chronotype PRS was associated with a higher overall LE8 score in UKB (β = 0.574; SE = 0.098; p = 3.82 × 10^-9^), but not in NHSII (β = 0.693; SE = 0.623; p = 0.27), after adjustment for the covariates used in the main MR analyses for each cohort.

**Table 3.**
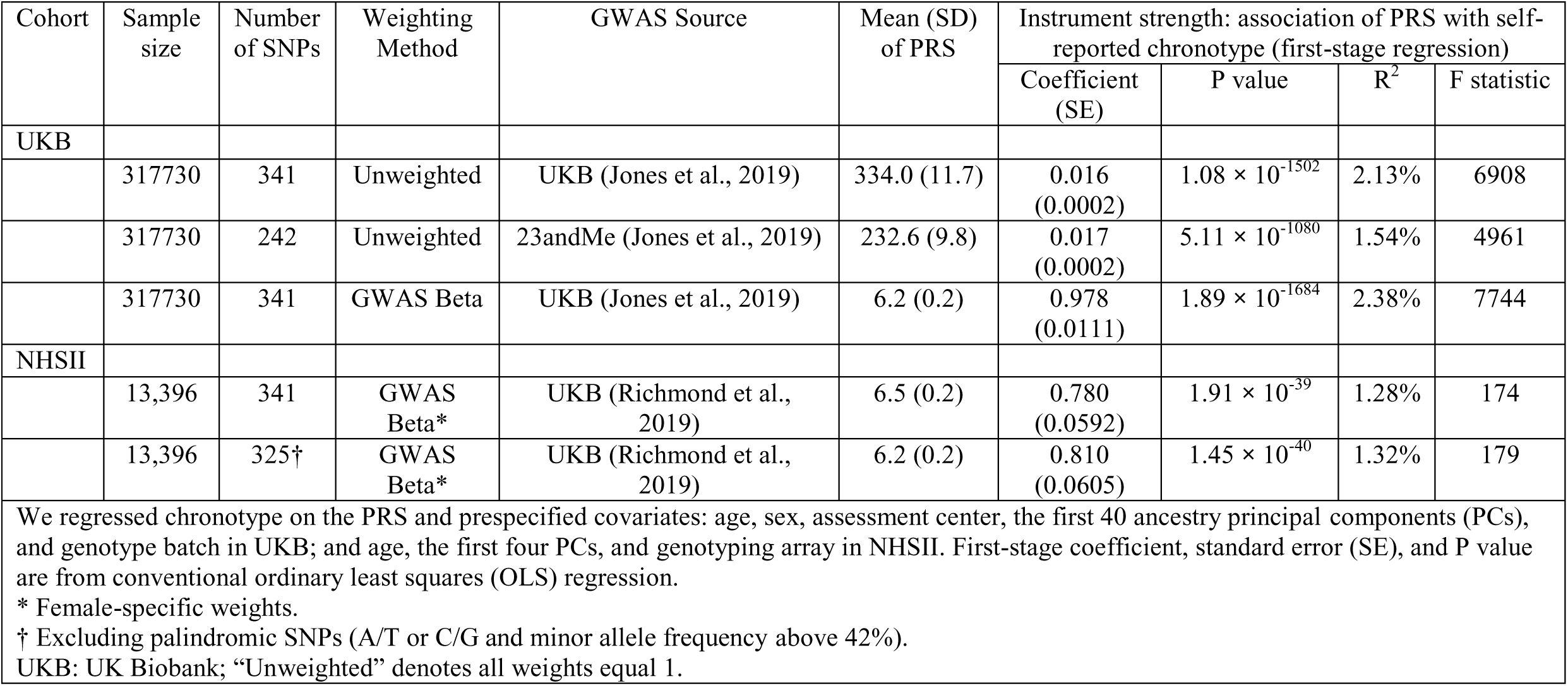
Polygenic Risk Score (PRS) for Chronotype in Analysis Cohorts.

In UKB, each one-unit increase in chronotype toward greater morningness (on the five-level measure) was associated with a 0.75-points higher overall LE8 score (95% CI: 0.55, 0.96; p < 9.26×10^-13^) (Figure 1). Although MR associations with individual LE8 components were all positive except body weight, only associations for diet (β = 2.70; 95% CI: 2.15, 3.26; p < 1.22×10^-21^), physical activity (β = 1.38; 95% CI: 0.87, 1.88; p < 9.85×10^-08^), and nicotine exposure (β = 0.77; 95% CI: 0.22, 1.32; p < 5.86×10^-03^) reached statistical significance (Table 4). Associations persisted after applying a Bonferroni correction. Results were similar when the PRS was derived by using the 242 SNPs replicated in the independent 23andMe GWAS or applying weights to the 341 SNPs (Supplemental Table 4). Results remained consistent in sensitivity analyses after excluding prevalent CVD (stroke/MI) cases (Supplemental Table 5), or after further adjustment for additional potential confounders (Supplemental Table 6).

**Table 4.** One-Sample Mendelian Randomization (2SLS) Estimates of the Associations Between Chronotype and Life’s Essential 8 (LE8) Scores, N = 317,730.

|  |  |  |  |  |
| --- | --- | --- | --- | --- |
|  | Genetic instrument: Chronotype Polygenic Risk Score |  |  |  |
|  | UK Biobank<br>341 unweighted SNPs from a UKB GWAS |  | Nurses' Health Study II<br>341 weighted SNPs from a female-specific UKB GWAS |  |
| LE8 scores<br>(range 0-100) | Beta (95% CI)* | P value | Beta (95% CI)† | P value |
| Diet | **2.70 (2.15, 3.26)** | **$1.22 \times 10^{-21}$** | **3.63 (0.44, 6.81)** | **0.0257** |
| Physical activity | **1.38 (0.87, 1.88)** | **$9.85 \times 10^{-08}$** | 1.17 (-2.32, 4.66) | 0.51 |
| Nicotine exposure | **0.77 (0.22, 1.32)** | **$5.86 \times 10^{-03}$** | 1.41 (-1.05, 3.86) | 0.26 |
| Sleep health | 0.18 (-0.15, 0.51) | $2.85 \times 10^{-01}$ | **1.95 (0.11, 3.79)** | **0.0380** |
| Body weight | -0.12 (-0.63, 0.39) | $6.47 \times 10^{-01}$ | -2.51 (-5.91, 0.89) | 0.15 |
| Blood lipid | 0.46 (-0.07, 0.99) | $8.77 \times 10^{-02}$ | NA | NA |
| Blood glucose | 0.32 (-0.03, 0.67) | $7.52 \times 10^{-02}$ | NA | NA |
| Blood pressure | 0.32 (-0.22, 0.86) | $2.42 \times 10^{-01}$ | -0.31 (-3.32, 2.70) | 0.84 |
| Overall score | **0.75 (0.55, 0.96)** | **$9.26 \times 10^{-13}$** | 0.89 (-0.67, 2.44) | 0.26 |
| Results from two-stage least squares Mendelian randomization analyses. Chronotype was modeled as a continuous variable ranging from -2 (definitely evening) to 2 (definitely morning). LE8 scores were continuous variables ranging from 0 to 100. Beta coefficients represent changes in LE8 score, in points, for each one-unit increase in the chronotype scale.<br>*Adjusted for age, sex, assessment center, first 40 ancestry principal components, and genotype batch.<br>†Adjusted for age, top four ancestry principal components, and genotyping arrays.<br>Blood lipid and blood glucose data were unavailable for most participants in the Nurses' Health Study II; therefore, these measures were excluded from the calculation of overall LE8 scores. NA: Not available in NHSII<br>Significance threshold after Bonferroni correction: $\alpha = 0.00625$ (0.05 / 8 tests). |  |  |  |  |

In the UKB, the associations for the overall LE8 score were similar in females (β = 0.79; 95% CI: 0.52, 1.07) and males (β = 0.71; 95% CI: 0.40, 1.02; Cochran’s Q test p: 0.70; Supplemental Table 7). Additionally, sensitivity analyses based on two-sample MR approaches

(e.g., weighted median) suggested limited evidence of horizontal pleiotropy. Estimates from IVW fixed and random-effects, weighted median, and weighted mode (phi = 2) methods were consistent with the primary 2SLS result; however, the weighted-mode estimate with a narrower bandwidth (phi = 1) was null and imprecise (suggesting a smaller number of SNPs clustered near no effect) and we observed heterogeneity in SNP effects (Cochran’s Q = 837, p <0.001; I^2^ = 59%; Figure 1, Supplemental Figure 2).

In NHSII, each one-unit increase in chronotype toward greater morningness was associated with a 0.89-points higher overall score (based on six measured LE8 components), though this association was imprecise and did not reach statistical significance (95% CI: -0.67, 2.44; p = 0.26) (Figure 1). However, associations were observed for two of the LE8 components, diet (β = 3.63; 95% CI: 0.44, 6.81; p = 0.0257) and sleep health (β = 1.95; 95% CI: 0.11, 3.79; p = 0.0380) (Table 4). Further, we observed significant associations when we limited the overall score to only the behavioral components, defined as the average of the diet, physical activity, nicotine exposure and sleep health scores (β = 2.04; 95% CI: 0.43, 3.65; p = 0.0130). Findings were consistent when palindromic SNPs were excluded from the weighted PRS (Supplemental Table 8) or after excluding participants with recent night shift work or prevalent CVD (Supplemental Table 9). Similar results were obtained when we used the DASH diet instead of AHEI-2010 in constructing the LE8 diet component (β = 3.29; 95% CI: 0.10, 6.48; p = 0.0436). However, none of the associations described above retained statistical significance after applying Bonferroni correction.

Lastly, meta-analysis yielded a significant association between increased morningness and higher overall LE8 score without evidence for between-study heterogeneity in the one-sample MR data (β = 0.75; 95% CI: 0.55, 0.96; p for Cochran’s Q = 0.86; Figure 1).

## DISCUSSION

In two large cohorts of middle-aged and older adults, we observed cross-sectional associations between self-reported greater morning chronotype and better cardiovascular health (LE8 score), after adjusting for a comprehensive list of sociodemographic, shift work, and other health-related characteristics as potential confounding factors. Results from individual-level one-sample MR analyses were consistent with the cross-sectional findings, demonstrating similar direction and magnitude of associations across both cohorts and in pooled analyses. However, in NHSII, the association was only observed when the overall score included behavioral components alone. Comparable associations were observed among both female and male participants in UKB. Greater morningness was associated with better diet, physical activity, and nicotine exposure scores in UKB, and with better diet and sleep health scores in NHSII. Sensitivity analyses largely corroborated the primary findings.

### Interpretation

Our findings provide evidence in favor of a causal relationship between greater morningness and better cardiovascular health profiles, as measured by the overall LE8 score. Most evidence to date comes from conventional observational studies focused on individual cardiovascular risk factors, which have reported associations between evening chronotype and suboptimal cardiovascular health (17). In our previous work, individuals with a definite evening chronotype were 54% more likely to have an overall unhealthy lifestyle in NHSII (N = 63,676), and 79% more likely to have a poor LE8 score in UKB (N = 322,777), compared to those with a definite morning chronotype (11, 12). Another cross-sectional study among 506 women also reported associations between evening chronotype and a low Life’s Simple 7 score, the predecessor of LE8 (34). No previous MR analysis has examined chronotype in relation to overall cardiovascular health.

We found greater morningness was associated with better scores in the four LE8 behavioral components: diet, physical activity, nicotine exposure, and sleep, with diet associations most consistent across cohorts, study designs, and all sensitivity analyses. By contrast, chronotype was associated with the four clinical factors (body weight, blood lipids, blood glucose, and blood pressure) only in cross-sectional analyses, but not in MR. Existing evidence on the associations between chronotype and individual CVD risk factors mostly comes from non-MR studies. Across three systematic reviews of 24 to 39 observational studies, evening chronotype (vs. morning) was associated with higher blood glucose, glycated hemoglobin, LDL cholesterol, and triglycerides; however, no associations were found for insulin, blood pressure, or total cholesterol and results were inconsistent for anthropometric factors and HDL cholesterol (16, 35, 36). Another nutrition-focused systematic review of 43 observational studies found that evening chronotype is associated with unhealthy eating habits, such as late-night eating and higher intake of ultra-processed foods, while morning chronotype have healthier eating behaviors (37).

A few MR studies have evaluated the associations between chronotype and some of the individual LE8 components. Similar to our findings, a recent two-sample MR analysis using data from 23andMe and UKB found that morning preference, compared to evening preference, was associated with greater consumption of foods linked to healthier diet quality (e.g., fresh fruits) and lower intake of unhealthy items (e.g., processed meats) (38). Additionally, consistent with our findings, MR studies have found no effect of chronotype on BMI or blood glucose (8, 39, 40). However, unlike our finding of no effect of chronotype on blood pressure, another two-sample MR study reported that greater morningness was associated with higher blood pressure; notably, that study used a subset of 57 SNPs from the 341 SNPs included in our analysis (41). Finally, an individual-level one-sample MR analysis found that higher cigarette smoking was associated with lower odds of morningness (using an instrument based on a single genetic variant associated with smoking); however, this association was not observed in their two-sample MR analysis or in the reverse direction (42).

Given that we observed causal links only between chronotype and behavioral cardiovascular risk factors, our findings suggest that evening chronotype (a circadian preference) is more strongly associated with cardiovascular health-related behaviors than with intermediate measures of cardiovascular disease. The circadian clock coordinates various cardiovascular functions, and disruption of its ∼24-hour rhythm has been linked to cardiometabolic disease (3). While LE8 captures diet quality, exercise amount, and sleep duration, the timing of these behaviors also plays an important role, although not assessed in this study; in evening chronotypes, circadian misalignment can disrupt sleep and meal timings, and limit opportunities for physical activity, thereby contributing to less favorable LE8 scores (3, 13). Interventions that aim to address this misalignment might help to improve cardiovascular health among evening chronotypes.

### Strengths and limitations

Strengths of our study include the use of two large well-established cohorts for comparison and replication, integration of cross-sectional and MR approaches (i.e., triangulation of evidence), and comprehensive assessment of CVD risk factors based on the LE8 framework. Further, we used one-sample MR, which compared with summary-statistics MR, offers strengths such as individual-level confounder adjustment, direct comparison of observational and MR findings within the same participants, and avoidance of issues like data harmonization and sample heterogeneity (19). The chronotype PRS was strongly associated with chronotype in both cohorts, meeting the MR relevance assumption. We used a comprehensively derived LE8 score for cardiovascular health, incorporated objective measures in UKB, and adjusted for comprehensive confounders, including detailed shift work history in NHSII.

Our study has some limitations. Participants were exclusively of White ethnicity, limiting generalizability to other ancestry groups. Selection and “healthy volunteer” bias are possible, particularly given UKB’s low response rate (∼6%) (20). However, many previously observed associations in UKB have been replicated in more representative studies of the general UK population or other populations, supporting the generalizability of our findings (43, 44). Both in our study and the discovery GWAS (8, 18), chronotype was assessed using a single self-reported MEQ item at a single time point, which may introduce measurement error; nonetheless, this item is highly correlated with the full MEQ score (25). Importantly, associations between the identified chronotype loci and accelerometer-derived sleep timing have been observed (8). Moreover, while chronotype can change over time, we observed robust within-individual stability in both cohorts, suggesting that a single-time measurement likely did not introduce substantial misclassification (11, 12). Further, we treated chronotype as a linear trait (as in the discovery GWAS), which may have missed potential nonlinear associations. Specifically, those at either extreme (e.g., extreme morning chronotype) may be at higher risk of circadian misalignment and poorer cardiovascular health. Lastly, chronotype may be an incomplete proxy for circadian misalignment. Alternative metrics that integrate self-reported and genetically determined chronotype with actual sleep timing may better capture circadian misalignment, which requires further investigation.

We used both cross-sectional and MR designs, each with inherent limitations. Cross-sectional findings are prone to measurement error, confounding, and reverse causation. MR can strengthen causal inference, though its own assumptions and limitations should be considered. We found associations between the chronotype PRS and some covariates (e.g., education). However, results remained consistent after adjusting for potential measured confounders, supporting the independence assumption. Moreover, because the overall LE8 score combines multiple components, the chance increases that chronotype SNPs may influence individual components through pathways other than chronotype (horizontal pleiotropy). However, MR estimates showed generally consistent association directions across all components, in line with the overall LE8 score. Additionally, we observed null and inconsistent estimates for the weighted mode, likely due to its low precision and reliance on the plurality-valid assumption (19). Sensitivity analysis with a wider bandwidth (phi = 2) yielded estimates consistent with other two-sample MR methods, 2SLS, and cross-sectional results, providing evidence that the exclusion restriction assumption is largely met. In UKB analyses, winner’s curse bias (i.e., overestimation of SNPs’ effect sizes) is possible since the discovery GWAS was also conducted in UKB, which can bias one-sample MR findings toward cross-sectional results (19). To address this, we used an unweighted chronotype PRS and an alternative PRS based on 242 SNPs replicated in an independent 23andMe GWAS and found consistent associations (8). Lastly, measurement tools were not fully harmonized across the cohorts, and two of the LE8 components were unavailable in NHSII. These limitations may introduce measurement error, although a prior study supported the validity of reduced scores (29).

### Conclusion

In two large UK and US cohorts of middle-aged and older adults, we found both observational and MR evidence supporting that greater morningness is associated with better cardiovascular health, particularly health behaviors. We also found consistent evidence linking morningness preference to healthier diet, and some evidence for associations with physical activity, nicotine exposure, and sleep. Our findings suggest that chronotype may be an important determinant for cardiovascular risk behaviors that could be integrated in risk assessment to inform more personalized prevention strategies. Further research is needed to clarify the potential nonlinear associations between chronotype and cardiovascular health and to elucidate the underlying mechanisms.

## Data Availability

This study used data from UK Biobank and the Nurses' Health Study II (NHSII). Individual-level participant data cannot be publicly shared due to participant consent, confidentiality protections, and data use agreements. UK Biobank data are available to qualified researchers through application to the UK Biobank resource. NHSII data are maintained by the Nurses' Health Studies (Channing Division of Network Medicine, Brigham and Women's Hospital and Harvard Medical School) and may be made available to qualified investigators upon reasonable request and with approval in accordance with the cohort's data sharing policies. Programs and analytic code used to generate the results can be shared by the first author upon request.

https://nurseshealthstudy.org/researchers

https://www.ukbiobank.ac.uk/enable-your-research

## Acknowledgements

The authors gratefully acknowledge the participants of the UK Biobank and Nurses’ Health Study II for their invaluable contributions of time and data, as well as the UK Biobank and the Channing Division of Network Medicine at Brigham and Women’s Hospital for their continued support of this work. The authors also acknowledge the funding sources that made this research possible.

